# An Intrinsic and Extrinsic Evaluation of Learned COVID-19 Concepts using Open-Source Word Embedding Sources

**DOI:** 10.1101/2020.12.29.20249005

**Authors:** Soham Parikh, Anahita Davoudi, Shun Yu, Carolina Giraldo, Emily Schriver, Danielle L. Mowery

**Affiliations:** School of Engineering and Applied Science, University of Pennsylvania; Department of Biostatistics, Epidemiology, & Informatics, University of Pennsylvania; Division of Hematology/Oncology, Department of Medicine, Penn Medicine; Data Analytics Center, Penn Medicine; Institute for Biomedical Informatics, University of Pennsylvania

## Abstract

**Introduction:** Scientists are developing new computational methods and prediction models to better clinically understand COVID-19 prevalence, treatment efficacy, and patient outcomes. These efforts could be improved by leveraging documented, COVID-19-related symptoms, findings, and disorders from clinical text sources in the electronic health record. Word embeddings can identify terms related to these clinical concepts from both the biomedical and non-biomedical domains and are being shared with the open-source community at large. However, it’s unclear how useful openly-available word embeddings are for developing lexicons for COVID-19-related concepts.

**Objective:** Given an initial lexicon of COVID-19-related terms, characterize the returned terms by similarity across various, open-source word embeddings and determine common semantic and syntactic patterns between the COVID-19 queried terms and returned terms specific to word embedding source.

**Materials and Methods:** We compared 7 openly-available word embedding sources. Using a series of COVID-19-related terms for associated symptoms, findings, and disorders, we conducted an inter-annotator agreement study to determine how accurately the most semantically similar returned terms could be classified according to semantic types by three annotators. We conducted a qualitative study of COVID-19 queried terms and their returned terms to identify useful patterns for constructing lexicons. We demonstrated the utility of applying such terms to discharge summaries by reporting the proportion of patients identified by concept for pneumonia, acute respiratory distress syndrome, and COVID-19 cohorts.

**Results:** We observed high, pairwise inter-annotator agreement (Cohen’s Kappa) for symptoms (0.86 to 0.99), findings (0.93 to 0.99), and disorders (0.93 to 0.99). Word embedding sources generated based on characters tend to return more lexical variants and synonyms; in contrast, embeddings based on tokens more often return a variety of semantic types. Word embedding sources queried using an adjective phrase compared to a single term (e.g., dry cough vs. cough; muscle pain vs. pain) are more likely to return qualifiers of the same semantic type (e.g., “dry” returns consistency qualifiers like “wet”, “runny”). Terms for fever, cough, shortness of breath, and hypoxia retrieved a higher proportion of patients than other clinical features. Terms for dry cough returned a higher proportion of COVID-19 patients than pneumonia and ARDS populations.

**Discussion:** Word embeddings are a valuable technology for learning terms, including synonyms. When leveraging openly-available word embedding sources, choices made for the construction of the word embeddings can significantly influence the phrases returned.

## Introduction

COVID-19 has become a devastating pandemic felt throughout the world. Scientists are developing new methods for determining infection rates, disease burden, treatment efficacy, and patient outcomes [1]. Our ability to distinguish COVID-19 patients from non-COVID-19 patients for clinical and translational studies requires clinical symptomatology, radiological imaging, laboratory tests, and associated disorders derived from electronic health record (EHR) data. Much of this information is locked within the EHR clinical notes. To accurately characterize each patient’s COVID-19 profile for study, we must develop natural language processing (NLP) systems to reliably extract COVID-19-related information. One of the first steps to extracting this information is developing lexicons with adequate coverage for all synonyms describing each COVID-19 concept. In the clinical domain, lexicons have been developed using several techniques: standardized vocabularies [2], lexico-syntactic patterns [3], term expansion [4], and distributional semantics [5]. Moreover, word embedding technologies have become increasingly popular for identifying semantically and syntactically-related terms within vector spaces by assessing the ***distributional hypothesis*** that “*words that share a common, relative vector space will often also share a common, semantic relatedness [5]*.”

### Word embeddings

Word embeddings represent a word in a vector space while preserving its contextualized usage. Word embeddings have been leveraged to learn synonyms to develop lexicons [6]. These vectors are commonly learned by training algorithms like Word2Vec [7], FastText [8] and GloVe [9] on large corpora of texts including domain-independent texts (e.g., internet web pages like Wikipedia and CommonCrawl; social media like Twitter and Reddit) and domain-specific texts (e.g., clinical notes like the **M**edical **I**nformation **M**art for **I**ntensive **C**are III (MIMIC III) database notes and biomedical research articles like PubMed). These domain-specific embeddings may capture richer biomedical information than open-domain embeddings (e.g., **Standard GloVe embeddings**). For example, **BioASQ** released their embeddings trained using the Word2Vec algorithm on 11 million biomedical abstracts from PubMed [10]. Moen and Ananiadou trained embeddings using Word2Vec on a combination of PubMed and PubMed Central articles along with Wikipedia in order to combine open-domain and biomedical knowledge (**BioNLP** corpus) [11]. Zhang et al. 2019 [12] (**BioWordVec** corpus) and Flamholz et al. 2019 (**ClinicalEmbeddings** corpus) [13] also leverage PubMed and PubMed Central articles in addition to clinical notes from the Medical Information Mart for Intensive Care (MIMIC III) to train embeddings using the FastText, GloVe, and Word2Vec algorithms [14].

### Word Embedding Evaluations

Systematic evaluations of word embeddings can be broadly classified into two categories, *intrinsic* and *extrinsic* evaluations. Intrinsic evaluations typically evaluate these word embeddings against human annotations by measuring the similarity or relationship between the queried and returned word pairs. Pakhomov et al. [15,16] and Pedersen et al. [17] have developed datasets containing pairs of biomedical terms along with their degree of relatedness as rated by human annotators. Furthermore, Pakhomov et al. [15] and Hliaoutakis et al. [18] have annotated pairs of medical terms for their semantic similarity. One intrinsic evaluation for these human annotations entails computing the Spearman’s coefficient between word pairs. Others have intrinsically evaluated word embeddings by clustering biomedical terms from the Unified Medical Language System (UMLS) and Ranker [19] and assessing the cluster quality using metrics like Davies-Bouldin Index and the Dunn Index. Word embeddings have advanced the state-of-the-art for many intrinsic, natural language processing (NLP) subtasks, i.e., reading comprehension [20], natural language inference [21], text summarization [22], vocabulary development [6], and document classification [23]. An extrinsic or summative evaluation of clinical word embeddings can involve evaluating the performance of machine learning models by using word embeddings to complete a biomedical research task or clinical operation such as patient phenotyping [24,25], patient fall prediction [23], and patient hospital readmission prediction [26].

### COVID-19 and Word Embeddings

In recent years, there has been extensive work to leverage biomedical and clinical texts to develop word embeddings [27]. For example, clinical word embeddings have been trained to identify drugs [28], substance abuse terms [6], and anatomical locations [13]. More recently, word embeddings have been used to understand the COVID-19 pandemic. For example, Schild et al. trained word2vec models for learning terms related to “virus” (“corona”, “covid”, “wuflu”, “coronovirus”, “coronavirus”) for understanding the emergence of sinophobic behavior on web communities like Twitter and 4chan’s /pol/ facing COVID-19 outbreaks [29]. Klein et al. 2020 applied pre-trained Bidirectional Encoder Representations from Transformers (BERT) to identify Twitter users with probable or possible COVID-19 infection using their self-reported Twitter messages and temporal-spatial information [30]. However, to our knowledge, there has been no intrinsic evaluation of openly-available word embeddings to identify COVID-19 terms related to symptoms, findings, and disorder concepts for encoding clinical notes.

Our long-term goal is to develop a COVID-19 information extraction system to support a variety of purposes, including clinical and translational research, observational studies, clinical trials, public health monitoring, and hospital capacity monitoring. Our short-term goal is to conduct an *intrinsic evaluation* to 1) qualitatively analyze and compare various openly-available, word embedding sources by categorizing the most similar words returned for symptoms, findings, and disorders related to COVID-19 and 2) identify common patterns between returned terms and their associated COVID-19 query terms to better understand which of these word embedding sources and their configurations could best support synonym discovery as well as an *extrinsic evaluation* to 1) apply these terms and their learned synonyms to the discharge summaries of patients with pneumonia, acute respiratory distress syndrome, and COVID-19 and 2) report the proportion of patients identified for each concept for each disorder cohort.

## Materials and Methods

In this University of Pennsylvania Institute Review Board-approved study, we conducted a literature review of open-source word embeddings. We identified 7 publicly-available sources and characterized each source according to training source, unit of processing, context window embedding technology, preprocessing, embedding technology used, returned units, embedding size, and vocabulary size (**Table 1**).

**Table 1.** Description of word embedding sources used.

| Name | Source (author [cite]) footnote to url for download | Training Source | Unit | Context window | Preprocess (reduce case, remove stopwords, term types) | Embedding technology (gensim, FastText, GloVe, BERT, ELMO, etc.) | Returned unit (1-3 ngrams) | Embedding size | Vocab Size |
| --- | --- | --- | --- | --- | --- | --- | --- | --- | --- |
| BioNLP Lab PubMed + PMC W2V | Paper Vectors [11] | PubMed/ Pubmed Central articles | Token | 5 | mixed case, no stop words, skip-grams | word2Vec | 1 ngram | 200 | ~4 billion tokens |
| BioNLP LabWiki + PubMed + PMC W2V | Paper Vectors [11] | Wikipedia, PubMed/ Pubmed Central articles | Token | 5 | mixed case, no stop words, skip-grams | word2Vec | 1 ngram | 200 | ~5.4 billion tokens |
| BioASQ | Report Vectors [10] | PubMed abstracts | Token | 5 | lowercase, no stop words, continuous bag of words | word2Vec | 1 ngram | 200 | ~1.7 billion tokens |
| Clinical Embeddings W2V300 | Paper GitHub Link [31] | PubMed/ PMC/ MIMIC III | Token | 7 | lowercase, include stop words, skip-grams | word2Vec | 1-3 ngrams | 300 | ~300k tokens |
| BioWordVec Extrinsic | Paper GitHub Link Vectors [12] | PubMed + MeSH | Character | 5 | lowercase, include stop words | FastText | 1-3 ngrams | 200 | ~2.3 billion tokens |
| BioWordVec Intrinsic | Paper GitHub Link Vectors [12] | PubMed + MeSH | Character | 20 | lowercase, include stop words | FastText | 1-3 ngrams | 200 | ~2.3 million tokens |
| Standard GloVe Embeddings | Paper Vectors [9] | Common Crawl | Token | 10 | mixed case | GloVe | 1 ngram | 300 | ~2.1 billion tokens |

### Constructing the reference standard

We generated a list of terms for COVID-19-related semantic categories of *symptoms* (“fever”, “high fever”, “cough”, “wet cough”, “dry cough”, “congestion”, “nasal congestion”, “pain”, “chest pain”, “muscle pain”, “shortness of breath”, “dyspnea”, “tachypnea”, “malaise”, “headache”, “sore throat”), *findings* (“hypoxia”, “opacities”, “bilateral opacities”, “infiltrates”, “lung infiltrates”), and *disorders* (“ARDS”, “respiratory distress”, “acute respiratory distress syndrome”, “pneumonia”) described in [1]. We queried each word embedding source detailed in **Table 1** using these COVID-19-related phrases and retrieved the top 20 phrases based on ranked cosine similarity (terms closest to 1.0 signifying high similarity). Three annotators (a biomedical informatician, a clinical general internist/informatician, and a second-year medical student) encoded each returned phrase with the following semantic class types:

- **Negation (black)**: a negation of the query term, e.g., “afebrile” is a negation of “fever”
- **Synonyms (green)**: a lexical variant of the query term with highly similar or synonymous meaning including misspellings and short forms, e.g., “ARDS” is a synonym for “Acute Respiratory Distress Syndrome”.
- **Symptom/signs (yellow)**: any symptom, observation, finding, or syndrome that is not a synonym of the query term, e.g., “fever” is a symptom returned by “cough”
- **Disease/disorders (blue)**: any disease, disorder or diagnosis that is not a synonym for the query term, e.g., “pneumonia” is a disorder returned by “dyspnea”
- **Hyponym (light red)**: a more specific semantic type of the query term, e.g., “ground-glass opacities” is a hyponym of “opacities”
- **Hypernym (dark red):** a broader semantic type of the query term, e.g., “cough” is a hypernym of “productive cough”
- **Qualifiers (teal):** any non-clinical temporal, spatial, quality, extent, size descriptor, e.g., “dry” is a qualifier for “cough”.
- **Anatomical location (orange):** any clinical anatomical or positional descriptor, e.g., “lower lobe” is an anatomical location.
- **Therapeutic (purple):** any medication, therapy, or procedure, e.g., “mechanical ventilation” is a therapeutic device.
- **Other (grey):** any semantic type that was not among the aforementioned or a non-clinical type, e.g., “traffic” returned for “congestion”.

### Assessing Inter-annotator Agreement

For each annotator pair, we computed the inter-annotator agreement (IAA) for the semantic class types for each queried term using Cohen’s kappa [32] using sklearn [33]. Specifically, for each queried phrase e.g., “fever”, each annotator encoded the semantic type of the returned candidate term compared to the queried term, e.g., “pyrexia” encoded as a synonym for “fever”. We report the overall IAA by category (symptom, finding, and disorder) and by queried term (“fever”, “dry cough”). We also depict semantic disagreements between each pair of annotators using heatmaps generated using matplotlib [34].

### Analyzing the Similarity between COVID-19 Queried and Returned Terms

We depict the broad range of terms returned across openly-available, word embedding sources. For each queried term, the returned term will maintain the same semantic type across word embedding sources, but might return a different cosine similarity or occur in only select sources. Therefore, for all unique returned terms within the top 20 ranked by cosine similarity, we visualized the returned term based on its frequency among the word embedding sources at any rank using word clouds generated with matplotlib. The size of the word is a weighted representation of how frequently the returned term occurred across the 7 word embedding source (score is bounded between 0.14 (observed within only 1 of 7 word embedding sources) and 1.0 (observed within all 7 word embedding sources). Additionally, we plotted the range of cosine similarities for each returned term.

### Assessing the Semantic Distribution Patterns for Returned Candidate Terms by Source

We determined the distribution of semantic classes among returned candidates for each queried term according to word embedding source. Our goal is to identify common semantic themes among the queried-returned term pairs that might be driven by the word embedding source construction. We performed a content analysis of each semantic category as well as terms with and without modifiers to identify additional association patterns (**Table 2**).

**Table 2.** Queried terms (**symptoms, findings**, and **disorders**) with and without modifiers

|  | Without modifier | With modifier |
| --- | --- | --- |
| **Symptoms** | “fever” | “high fever” |
|  | “cough” | “wet cough”, “dry cough” |
|  | “congestion” | “nasal congestion” |
|  | “pain” | “chest pain”, “muscle pain” |
| **Findings** | “opacities” | “bilateral opacities” |
|  | “infiltrates” | “lung infiltrates” |
| **Disorders** | “ARDS” | “respiratory distress”,<br>“acute respiratory distress syndrome”, |

### Generating symptom severity profiles for pneumonia, ARDS, and COVID-19 patients

As a proof-of-concept, we compared the proportion of patients that can be classified according to COVID-19 illness severity groups using terms indicative of their clinical features for three cohorts: pneumonia, ARDS, and COVID-19 patients. For the pneumonia and ARDS patient cohort, we queried all inpatient encounters and the resulting discharge summaries with COVID-19-related disorders: ARDS (ICD codes: 518.5, 518.81, 518.82) and pneumonia (ICD codes: 480–488) from the MIMIC III database [14]. For the COVID-19 patient cohort, we queried all COVID-19 inpatient encounters from our EPIC PennChart COVID-19 registry and the resulting discharge summaries. In **Table 3**, we denote the clinical findings associated with COVID-19 respiratory illness severity categories [1]. We applied the expanded lexicon for COVID-19 respiratory illness severity clinical features using synonyms detected from all embedding approaches (*keywords + embedding expansion*). For each cohort, we report the proportion of patients with the clinical feature documented within one or more discharge summaries.

**Table 3:** Clinical Findings according to COVID-19 Respiratory Illness Severity groups

| COVID-19 Respiratory Illness Severity | Clinical Features |
| --- | --- |
| *Mild Illness* | mild fever, cough (dry), sore throat, malaise, headache, muscle pain, nasal congestion |
| *Moderate Pneumonia* | cough and shortness of breath |
| *Severe Pneumonia/*<br>*Acute Respiratory Distress Syndrome* | fever is associated with severe dyspnea, respiratory distress, tachypnea, and hypoxia |

## Results

We queried 7 embedding sources with 15 symptom terms, 5 finding terms, and 4 disorder terms resulting in 10,080 annotations (top 20 returned candidate terms x 25 queried terms x 7 word embedding sources x 3 annotators).

### Assessing Inter-annotator Agreement

We observed high overall pairwise inter-annotator agreement (IAA) between annotators for each semantic category: symptoms (0.86 to 0.99), findings (0.93 to 0.99), and disorders (0.93 to 0.99). For A1/A2 and A2/A3, we observed low to moderate IAA for “malaise” (0.40-0.41), “muscle pain” (0.6), “headache” (0.65-0.68), and “dry cough” (0.68). For A3/A1, IAA was consistently high (equal to or greater than 0.93). In **Figure 1**, we report the distribution of each queried term’s overall agreement between paired annotators. The color bar represents the third annotator pair.

**Figure 1.**
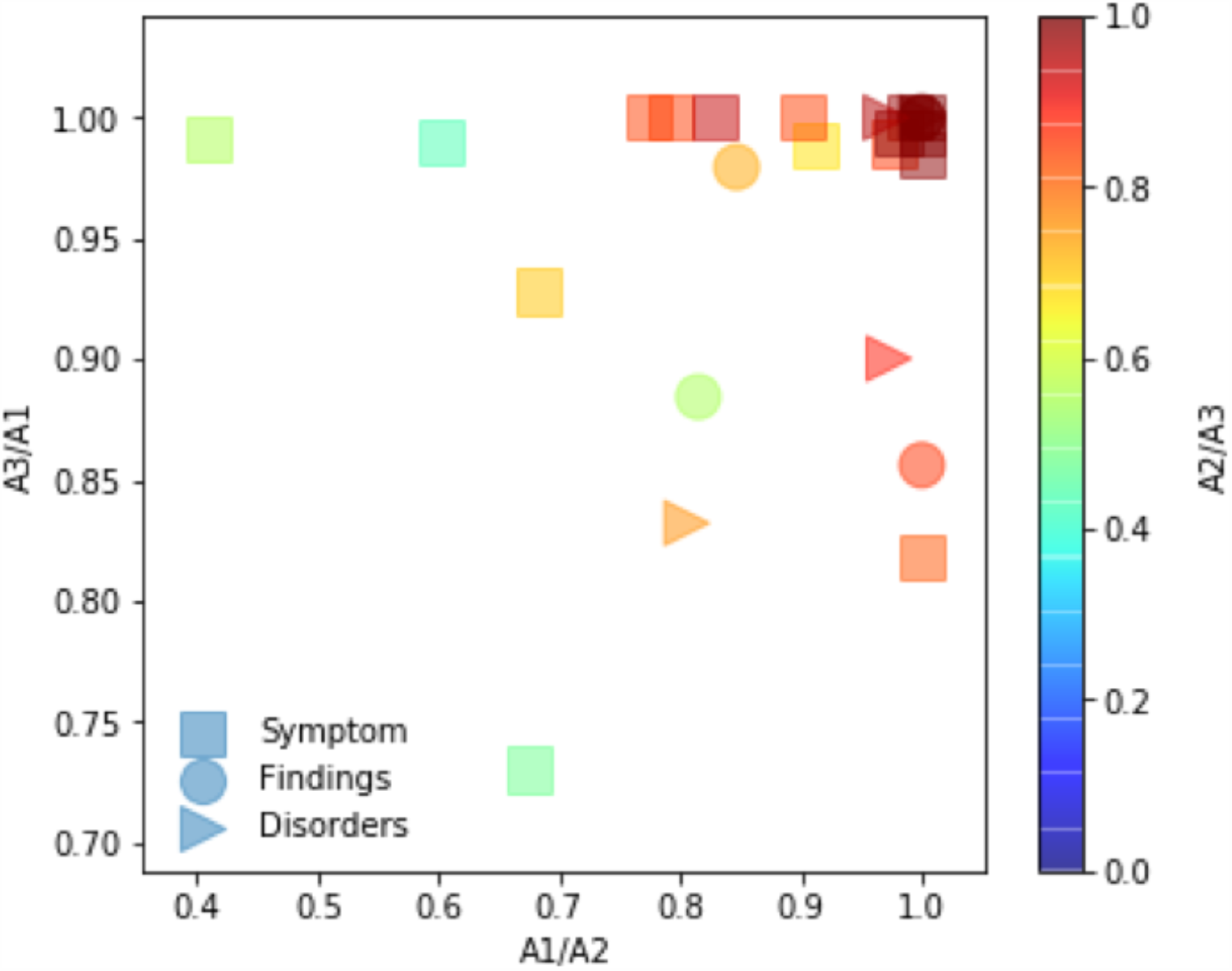
Pairwise inter-annotator agreement according to semantic category for each queried term. Semantic category types are represented as follows: square = symptoms/signs; circle = findings; triangle = disorders.

In **Figures 2-4**, for each returned term, we also computed IAA across semantic types. Across annotator pairs, we observed high IAA for all semantic types. Each heat map depicts systematic differences between annotators. In **Figure 2**, A1/A2 more often disagreed about whether a returned term was a more or less specific type or negation. In **Figure 3**, A2/A3 more often disagreed about whether a returned term was a synonym, disease/disorder, more or less specific type, other, or negated term. In **Figure 4**, A3/A1 most often disagreed about whether a returned term was a negated or other term.

**Figure 2.**
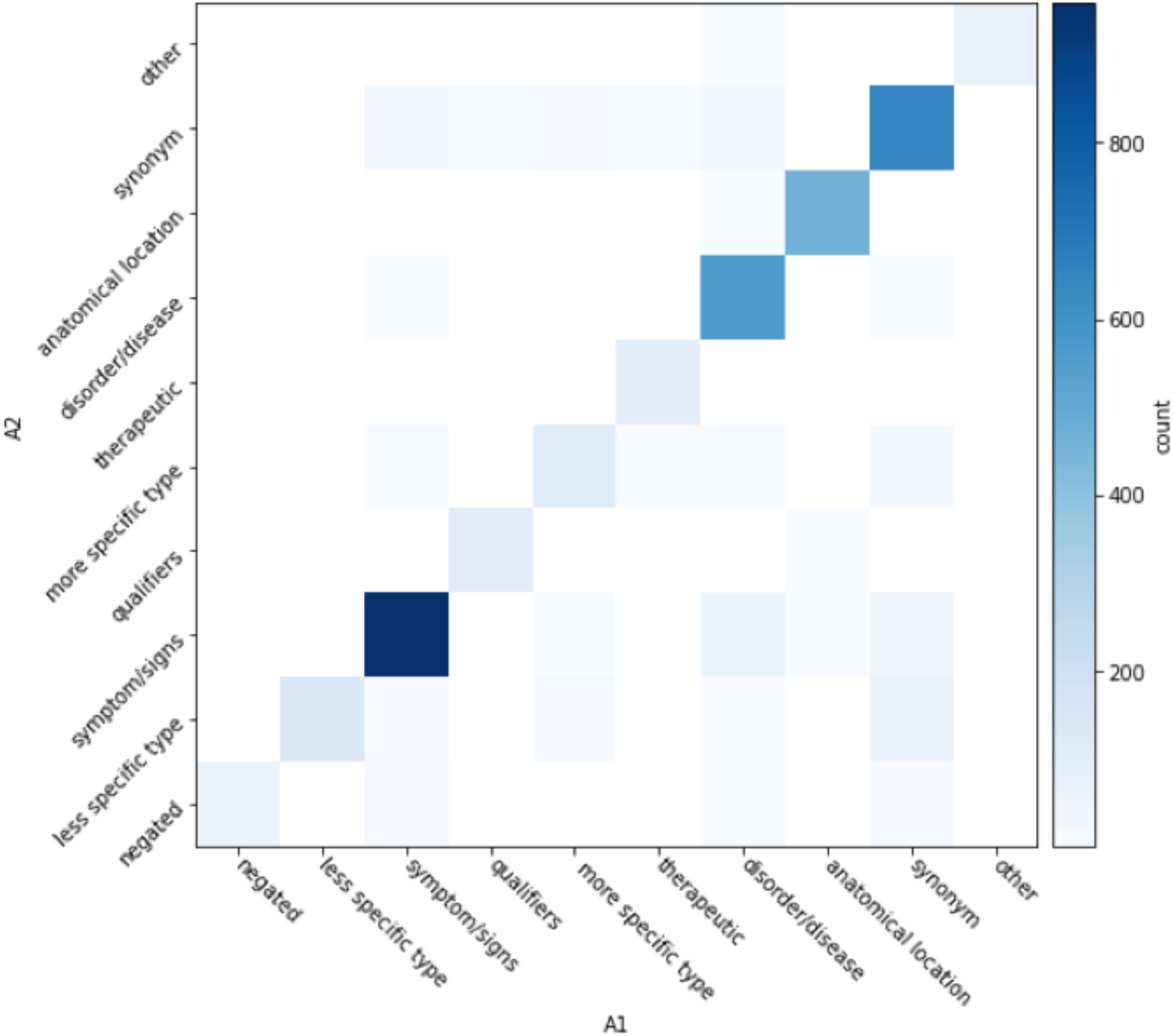
A1/A2 Inter-annotator agreement of returned terms according to semantic type.

**Figure 3.**
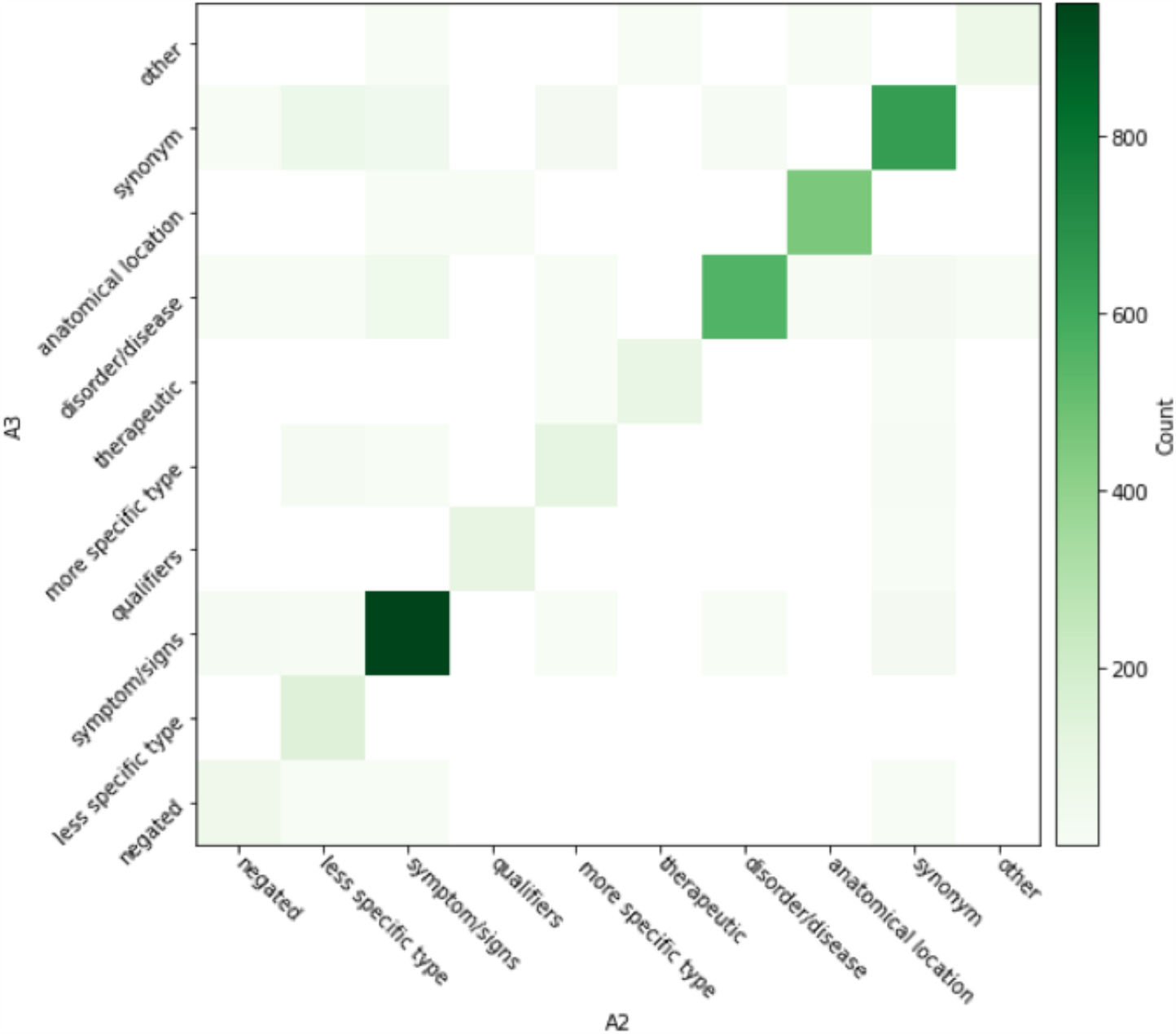
A2/A3 Inter-annotator agreement of returned terms according to semantic type.

**Figure 4.**
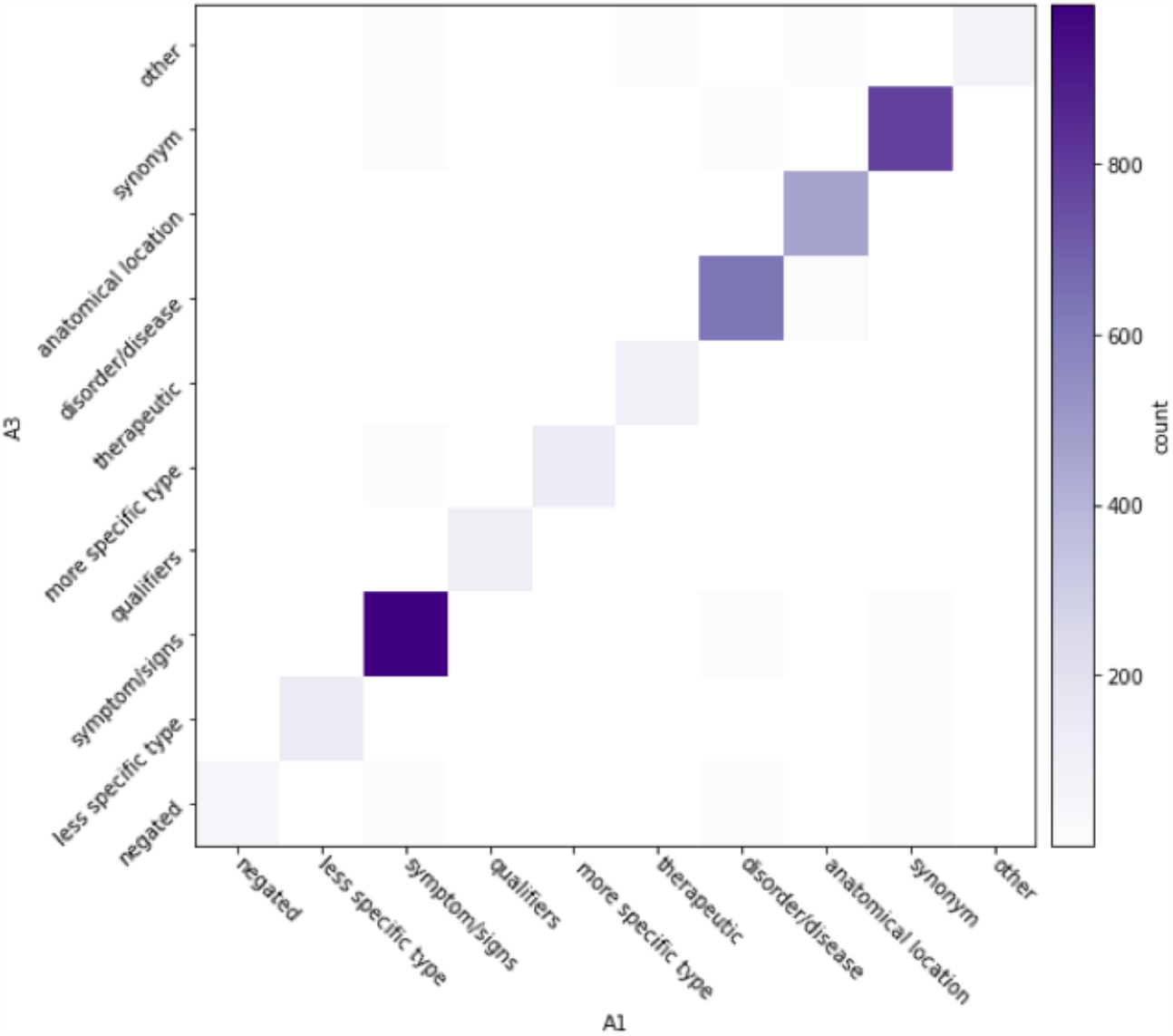
A1/A3 Inter-annotator agreement of returned terms according to semantic type.

### Analyzing the Similarity for Returned Candidate Terms

We report the broad range of queried terms returned across word embedding sources. For brevity, we depict three COVID-19-related concepts, one of each semantic category: *symptom* (“fever”; **Figure 5**), *finding* (“lung infiltrates”; **Figure 6**), and *disorder* (“acute respiratory distress syndrome”; **Figure 7**). For “fever”, synonyms, e.g., “pyrexia”, “fevers”, “febrile” and signs/symptoms, e.g., “chills”, “diarrhea”, were common among the returned terms. For “lung infiltrates”, the most frequent semantic types included anatomical locations, e.g., “lungs”, “peribronchial” and hypernyms, e.g., “infiltrate”, “infiltration” were among the returned terms. For “ARDS”, disease/disorders, e.g., “SARS”, “aSARS-CoV”, synonyms, e.g., “ards”, “respiratory-distress-syndrome” and hypernyms, e.g., “syndromee”, “syndrome-critical” were observed commonly among the returned terms.

**Figure 5.**
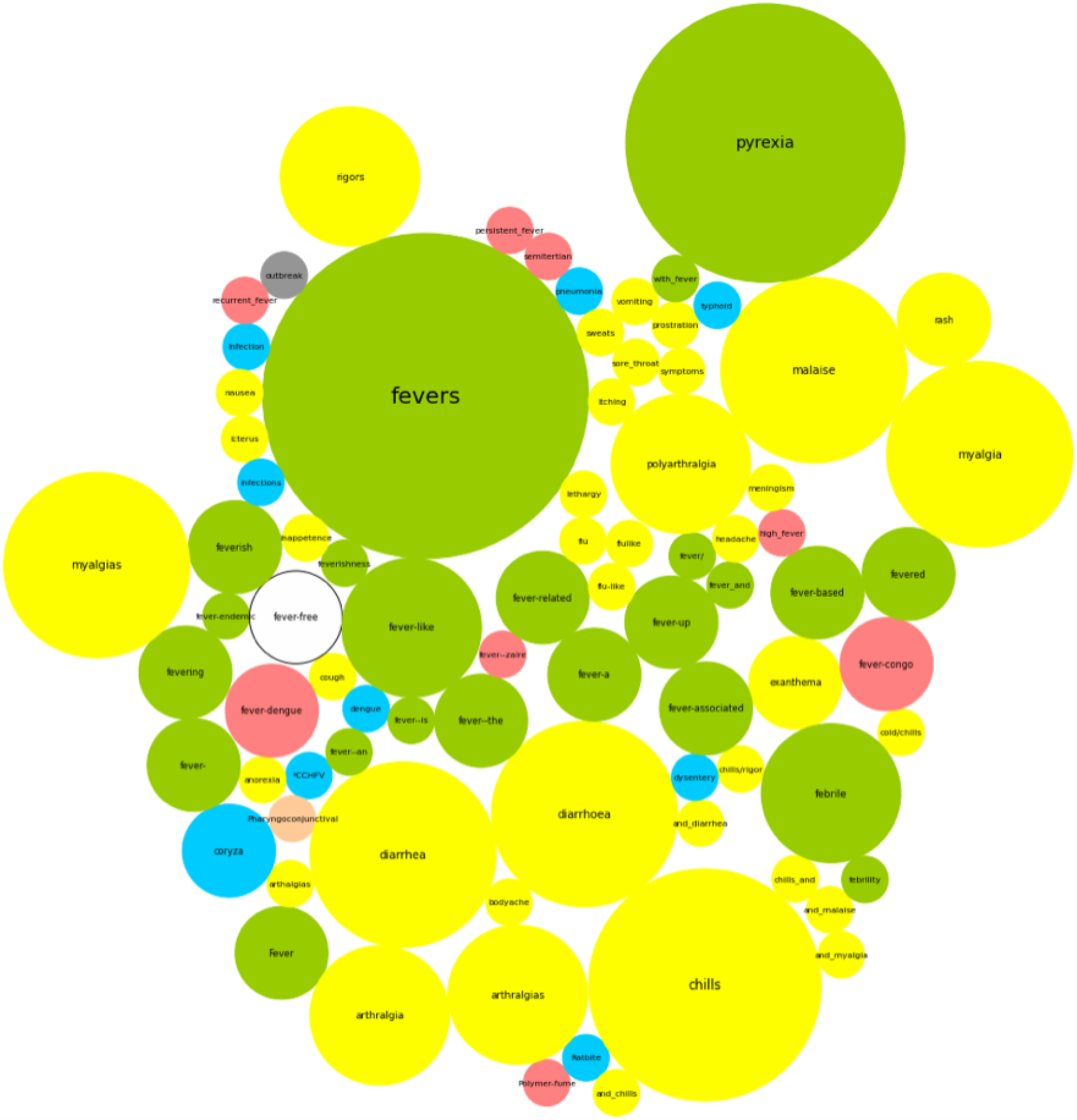
Word cloud depicting each returned term for “fever”. Colors correspond to semantic class types.

**Figure 6.**
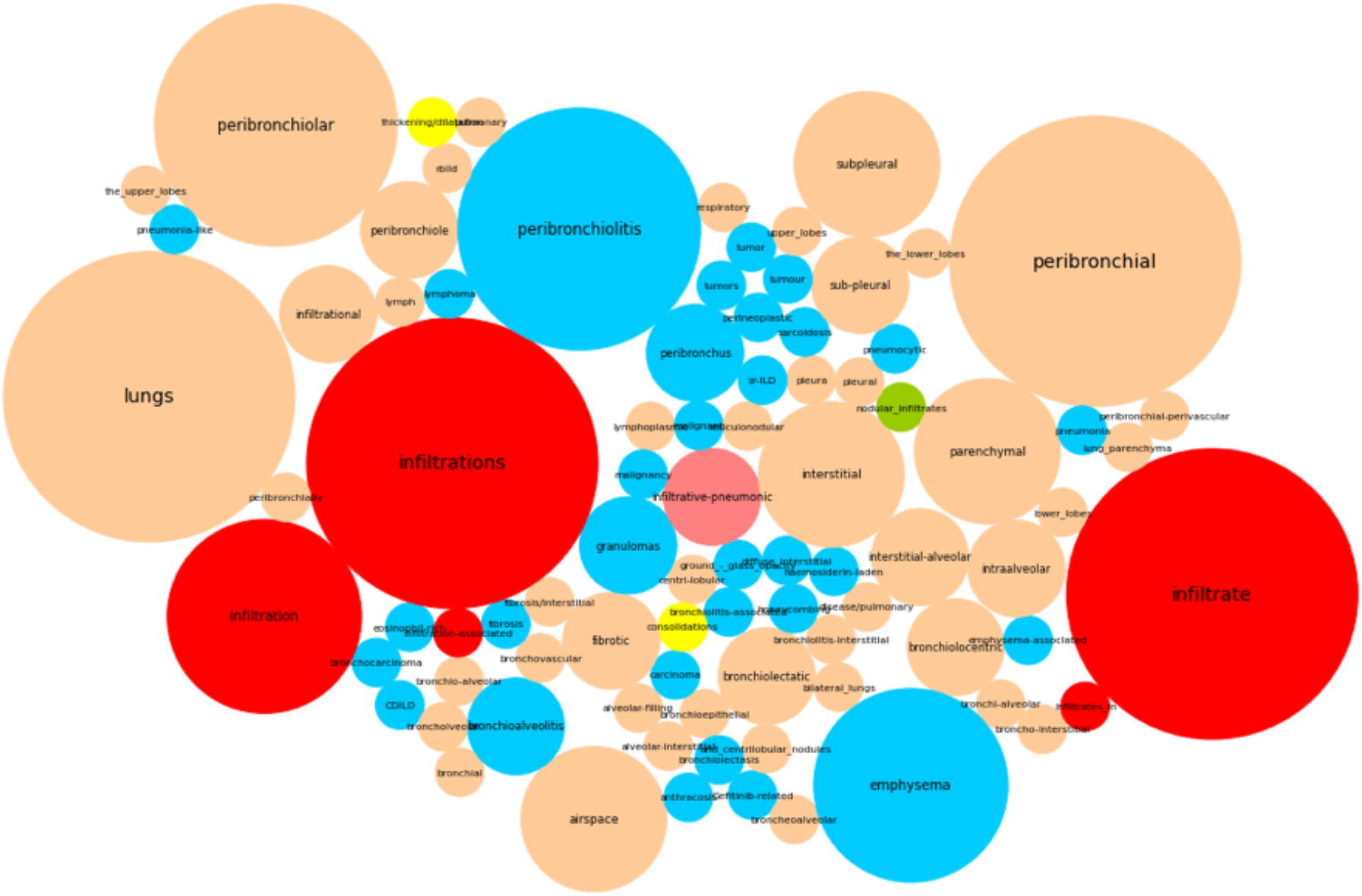
Word cloud depicting each returned term for “lung infiltrates”. Colors correspond to semantic class types.

**Figure 7.**
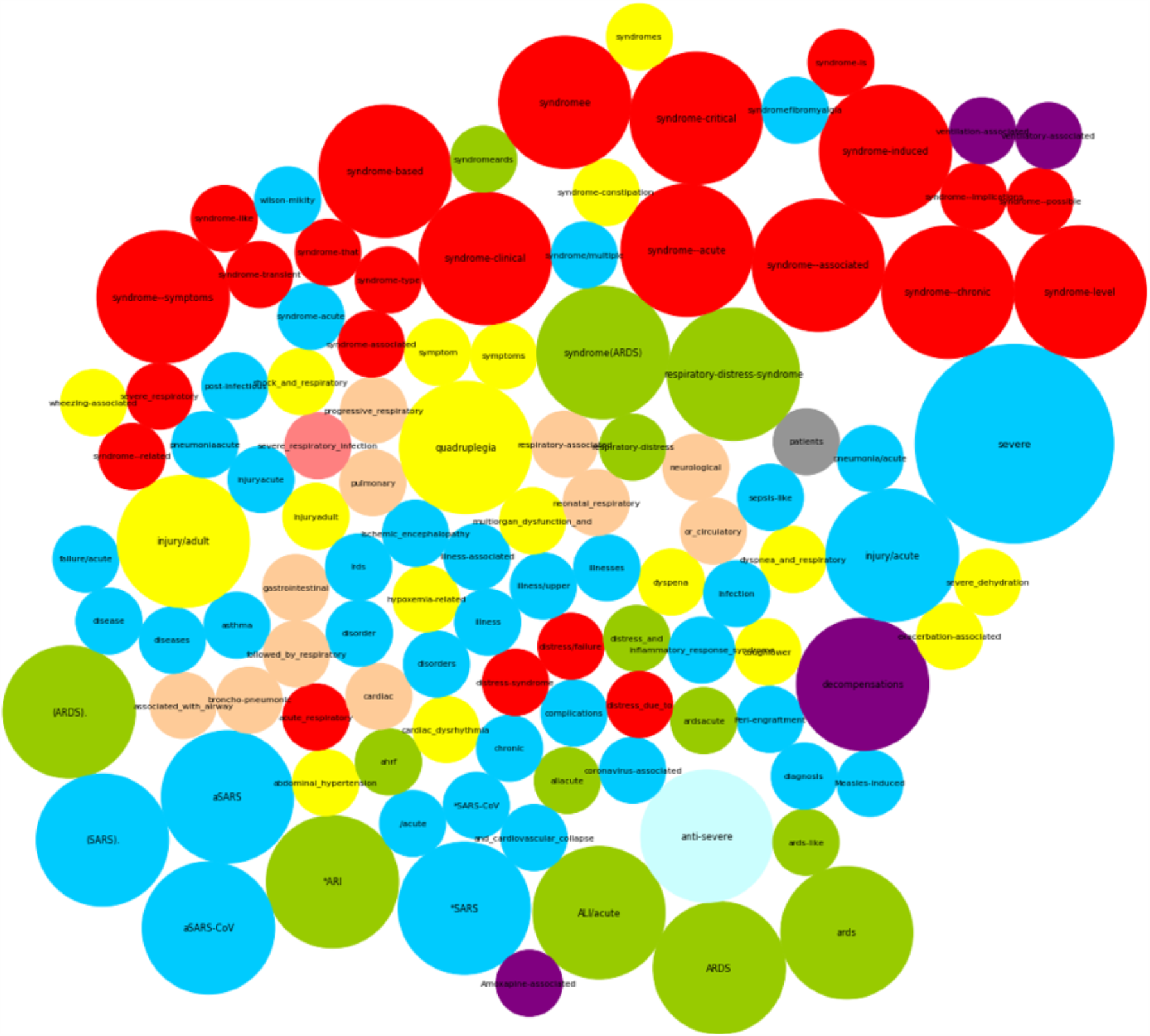
Word cloud depicting each returned term for “acute respiratory distress syndrome”. Colors correspond to semantic class types.

In **Figure 8**, we observe that given a queried term e.g., “fever”, “lung infiltrates”, and “acute respiratory distress syndrome”, returned terms differ by cosine similarity and variance. For example, some returned terms have high cosine similarity and low variability (left most in red and orange only); while others demonstrate variable cosine similarity and high variability (right most in all colors). Example queried: returned terms with *high cosine similarity and low variability* include: “fever”: “fevers”, “fevering”, “pyrexia”; “lung infiltrates”: “infiltration”, “infiltrates”, “peribronchial”; and “acute respiratory distress syndrome”: “syndrome(ARDS)”, “aSARS”, “syndromeards”. Example queried: returned terms with *variable cosine similarity and high variability* include: “fever”: “fevered”, “fever-based”, “fever-like”, “lung infiltrates”: “infiltrational”, “consolidations”, “bronchioepithelial”; and “acute respiratory distress syndrome”: “syndrome-is”, “syndrome-level”.

**Figure 8.**
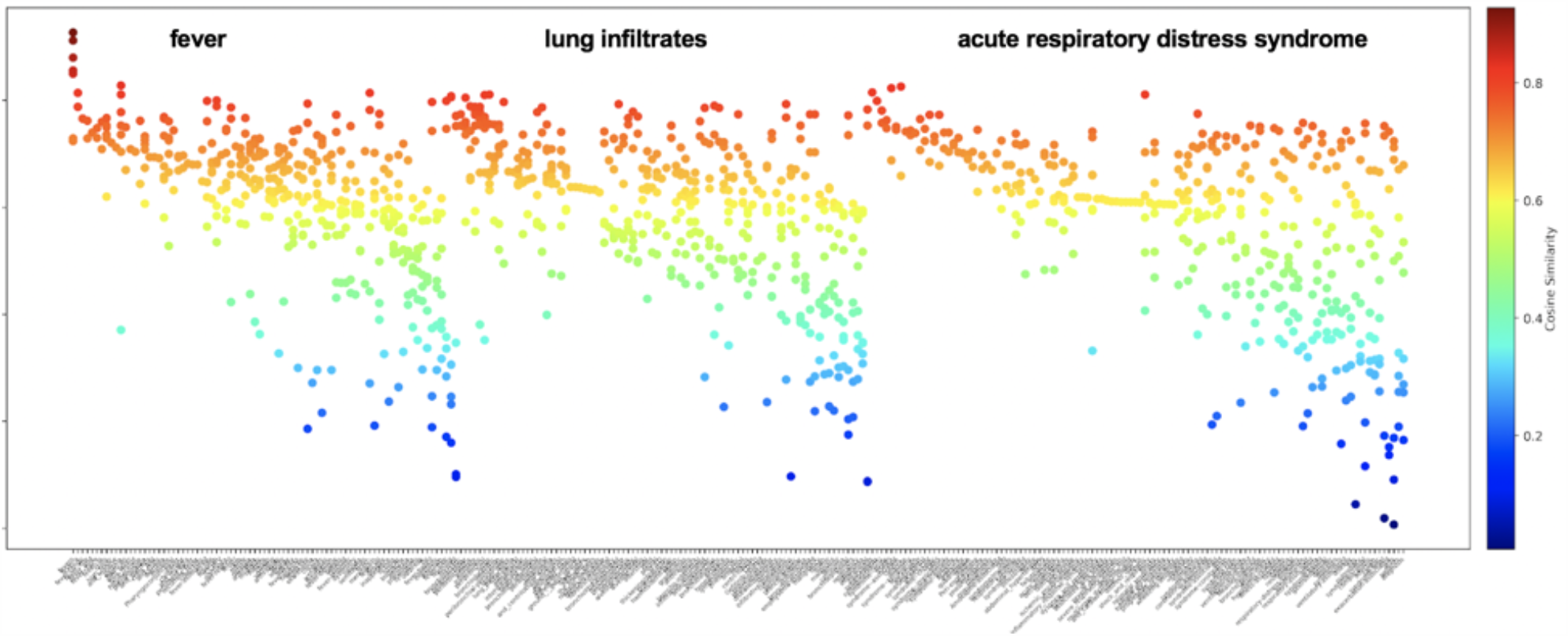
Cosine Similarity measures for each unique returned term among top 20 terms across all word embedding sources returned for the queried terms “fever”, “lung infiltrates”, and “acute respiratory distress syndrome”. Color range indicative of cosine similarity level (0.0-1.0), not semantic type.

### Assessing the Distribution of Semantic Types for Returned Candidate Terms by Source

We determined the distribution of semantic classes among returned candidates for each queried term according to word embedding source. Our goal is to identify common semantic themes among the queried and returned candidate term pairs that might be driven by the word embedding source construction. We observe that the **BioWordVec Extrinsic** and **BioWordVec Intrinsic** embeddings (**Figure 9e-f**) were more likely to generate synonyms (green), which is notably depicted for “fever”, “headache”, “hypoxia”, “dyspnea”, and “infiltrates”. We also observe more negation terms for “congestion”, “dry cough”, “pneumonia”. We observed less specific terms for “dry cough”, “wet cough”, and “acute respiratory distress syndrome”. Across the other word embeddings (**Figure 9a-d, g**), if a symptom/sign queried term was provided, we often observed a symptom/sign returned term. This also held true for disorders. For most word embedding sources (**Figure 9a-g**), we observed *qualifiers* were often returned when the queried term contained a *qualifier*, e.g., “dry cough” returns time and consistency qualifiers like “wet”, “runny”, etc. Furthermore, if a queried term contained an *anatomical location as an adjective* in the term phrase, e.g., “*nasal* congestion”, “*chest* pain”, and *“lung* infiltrates”, the returned terms were often *anatomical locations*. In few cases, the **Standard GloVe embeddings, BioWordVec Extrinsic**, and **BioWordVec Intrinsic** embeddings returned some terms with common term usage, e.g., “congestion” returns “traffic”, “bypass” or stop words, e.g., “and”, “a”, “of”.

**Figure 9:**
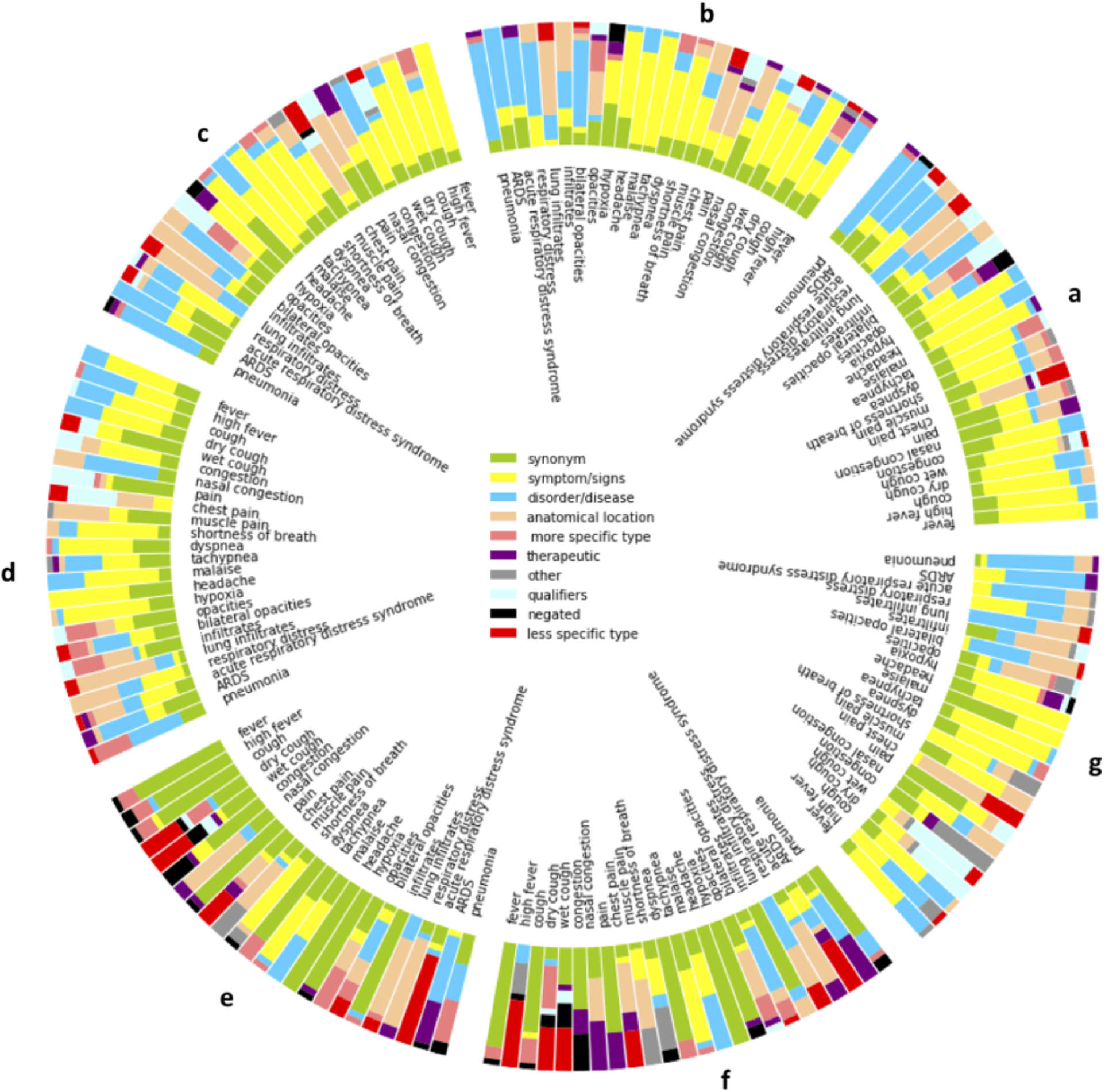
For each symptom, finding, and disorder queried term, the distribution of semantic types for returned term colored by semantic type for each embedding source: **(a)** BioNLP Lab PubMed + PMC W2V, **(b)** BioNLP LabWiki + PubMed + PMC W2V, **(c)** BioASQ, **(d)** Clinical Embeddings W2V300, **(e)** BioWordVec Extrinsic, **(f)** BioWordVec Intrinsic, and **(g)** Standard GloVe Embeddings.

### Generating symptom severity profiles for pneumonia, ARDS, and COVID-19 patients

**Figure 10** shows the proportion of patients from each disorder cohort (pneumonia, ARDS, COVID-19) that have one or more terms documented within their discharge summary representing clinical features from **Table 3**. The total number of patients in each cohort varied: pneumonia (n=6410 patients), ARDS (n=8647 patients), and COVID-19 (n=2397 patients). Terms indicative of clinical features for fever, cough, shortness of breath, and hypoxia retrieved a higher proportion of patients than other clinical features. Terms indicative of dry cough returned a higher proportion of COVID-19 patients than from pneumonia and ARDS cohorts.

**Figure 10:**
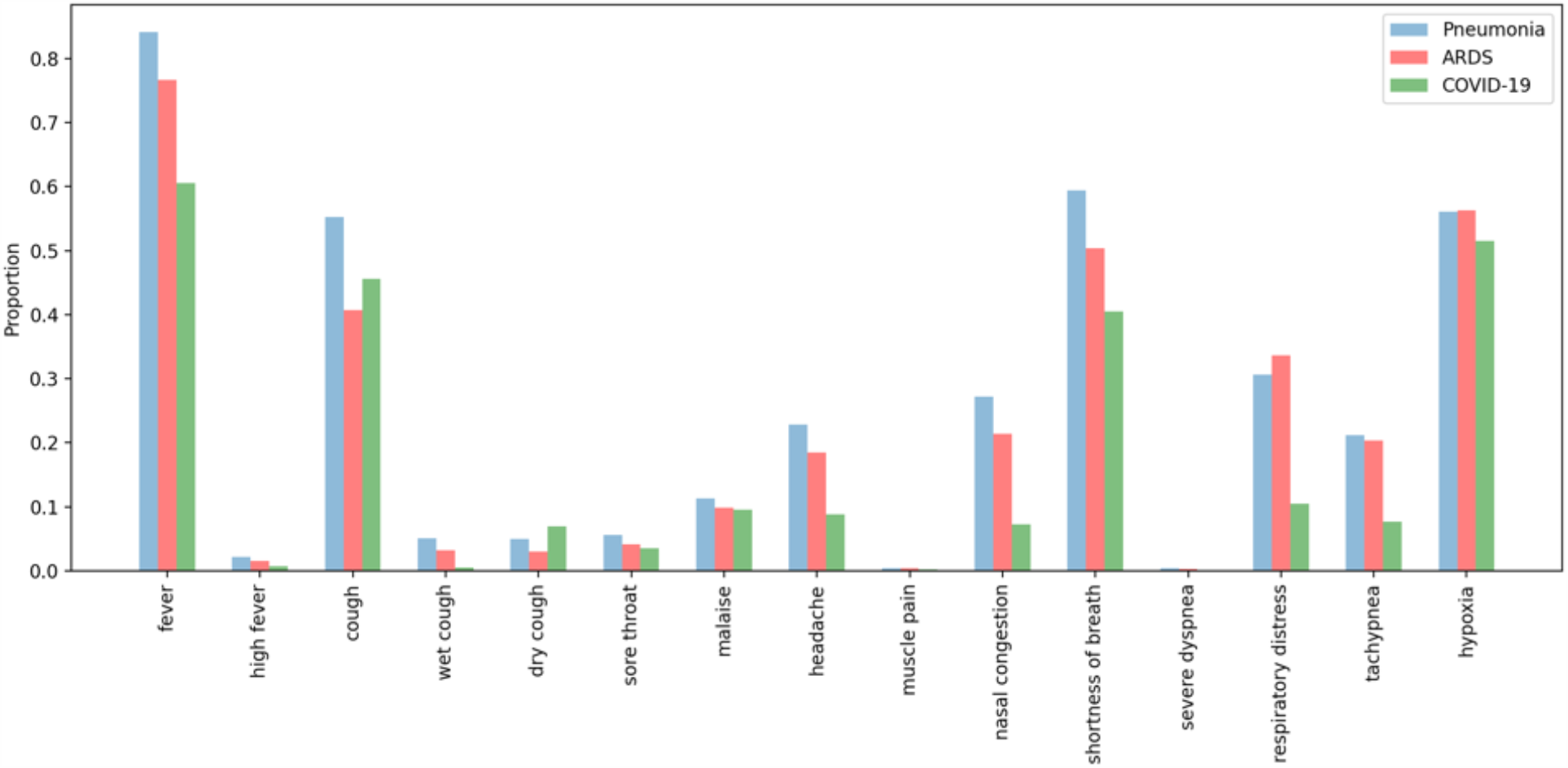
The proportion of patients with each clinical feature from **Table 3** documented within their discharge summary according to disorders (pneumonia, ARDS, and COVID-19).

## Discussion

### Assessing Inter-annotator Agreement

We observed high overall, pairwise inter-annotator agreement for symptoms, findings, and disorder categories. Annotators A1 and A3 were more often in agreement. For A1/A2 and A2/A3 pairs, we observed low to moderate IAA for queried terms such as “malaise”, “muscle pain”, “headache”, and “dry cough”. Annotators A1 and A3 systematically classified notably fewer returned terms as hypernyms and hyponyms than A2. For example, “migraine” is a hypernym for “headache”. Additionally, A2 more easily identified negated terms through medical terminology. Many cases required more clinical domain knowledge to make these distinctions, which were easier for the general internist.

### Analyzing the Similarity between COVID19 Queried and Returned Terms

When analyzing the cosine similarities between queried terms and returned terms, we observed that returned terms range from *high cosine similarity and low variability* to *variable cosine similarity and high variability*. We hypothesize that terms with high cosine similarity and low variability are more likely to be synonyms and useful for training an information extraction. In practice, the presence and cosine similarities of a term varied across word embedding sources. Our ability to identify and rank likely synonyms for lexicon development may be improved with additional processing steps and comparisons between the queried and returned terms for lexical similarity [35], morphological derivation [6], and short form construction/expansion [36].

### Assessing the Semantic Distribution Patterns for Returned Candidate Terms by Source

We determined the distribution of semantic classes among returned candidates for each queried term according to word embedding source. Our study intentions were to assess the distributional hypothesis that words with similar meanings are often used in similar contexts. Generally, if a symptom/sign queried term was provided, we often observed a symptom/sign returned term. This also held true for disorders. Furthermore, our goal was to identify common semantic themes among the queried and returned candidate term pairs that might be driven by the word embedding source construction. We observe that the **BioWordVec Extrinsic** and **BioWordVec Intrinsic** embeddings were more likely to generate synonyms. We hypothesize that this is likely due to training based on the characters rather than token; thus, the returned terms often share a common set of characters (queried term = “<u>fever</u>”; returned term = “<u>fever</u>ish”). Character-based embeddings will often return lexical variations of the queried term. Although **BioNLP, BipASQ**, and **Clinical Embeddings** generated fewer synonyms, these were often medical terms for the lay queried term (e.g., “lethargy” for “malaise”; “cephalea” for “headache”, “rhinorrhea” for “nasal congestion”). To maximize the diversity of learned synonyms, multiple approaches could be most beneficial. Returned negated terms were expressed with prefixes (e.g., “non-pneumonia-related”), suffixes (e.g., “fever-free”), or medical terminology (e.g., “normoxia”). Hypernyms were commonly observed among queried terms with an adjectival phrase (e.g., “high fever”, “muscle pain”, “dry cough”, “lung infiltrates”, etc.). Moreover, we observed qualifiers were often returned when the queried term contained a qualifier, e.g., time, consistency, and anatomical location qualifiers. For developing a clinical information extraction system, these returned terms can be useful for brainstorming synonyms as inclusionary terms as well as antonyms as exclusionary terms. We suspect that a mix of hypernyms and qualifiers were often returned, given the semantics of the individual parts of the queried phrase. It was not surprising that **Standard GloVe** embeddings returned some terms with common term usage, e.g., “congestion” returns “traffic”, “bypass” because they were trained using the CommonCrawl domain-independent corpora. Similarly, **BioWordVec Extrinsic** and **BioWordVec Intrinsic** occasionally return stop words as these were not removed prior to training and perhaps should be for detecting meaningful synonyms.

### Generating symptom severity profiles for pneumonia, ARDS, and COVID-19 patients

We created an expanded lexicon of COVID-19 respiratory illness clinical features (**Table 3**) using synonyms detected from all embedding approaches. We assessed the proportion of patients from three disorder cohorts (pneumonia, ARDS, COVID-19) with each clinical feature documented within their discharge summary. We observed that terms indicative of clinical features for fever, cough, shortness of breath, and hypoxia retrieved a higher proportion of patients than clinical features. For fever and cough, our lexicons for capturing contextualized mentions of these clinical features (e.g., high fever; wet or dry cough) retrieved modest proportions of patient cases. This is likely due to the variability of qualitative and quantifications of these symptoms (e.g., productive cough; fever 102 F) in discharge summaries. Terms indicative of dry cough returned a higher proportion of COVID-19 patients than pneumonia and ARDS populations. This is not surprising given this is a prominent symptom reported among COVID-19 patients.

### Limitations and Future Work

Our study has a few notable limitations. We began this study during the early stages of the COVID-19 pandemic when the symptomatology was less understood. COVID-19 is a heterogeneous disease with emerging symptomatology identified through ongoing clinical observational studies. Emerging COVID-19-related symptomatology, i.e., loss of smell and loss of taste and COVID toes were not included in our analysis as their association with COVID-19 were not well-understood at the time of our study. We leveraged existing word embedding sources to better understand the utility of embeddings for synonym generation. We recognize that further experimentation is needed to support broader claims of their utility. As a proof-of-concept of patient information retrieval, we applied an expanded lexicon of terms representing clinical features of COVID-19 to three disorder cohorts (pneumonia, ARDS, and COVID-19). Although these terms retrieved a high proportion of patients, we acknowledge that additional terms might be necessary to accurately identify these features and that contextualization (i.e., negation, severity, experiencer, temporality [37]) is critical to generating accurate patient profiles. We look forward to addressing these issues as next steps toward developing our clinical information extraction pipeline in our follow-up study.

## Conclusion

Word embeddings are a valuable technology for learning semantically and syntactically-related terms including synonyms, useful for text classification, information extraction, among other NLP tasks. When leveraging openly-available word embedding sources, choices made in the development of the embeddings can significantly influence the types of phrases and information returned.

## Data Availability

The annotated terms will be available through Github in January of 2021.

## Acknowledgements

We extend our gratitude to the open-source community for making their embeddings and data publicly available.

## Contributions

DM designed and conceptualized the study. ES wrangled Penn Medicine clinical notes for COVID-19 patients; AD wrangled MIMIC III data for pneumonia patients. SY, CG, and DM annotated terms. SP and AD conducted analyses. AD created all visualizations. All authors contributed to writing the manuscript and approved the final version.

## Informed Consent/IRB Statement

This study was reviewed and approved by the University of Pennsylvania Institutional Review Board (#831895,#843620).

## Abbreviations

COVID-19: Coronavirus Disease of 2019
NLP: Natural Language Processing
MIMIC III: Medical Information Mart for Intensive Care version 3
UMLS: Unified Medical Language System
IAA: Inter-Annotator Agreement

## Supplemental Materials

**Table 1.** Overall agreement by COVID19 category and by queried term for each annotator pair

| Category | Queried Term | Annotator1/<br>Annotator2 | Annotator2/<br>Annotator3 | Annotator3/<br>Annotator1 |
| --- | --- | --- | --- | --- |
| **Symptoms** |  | 0.87 | 0.86 | 0.99 |
|  | fever | 0.91 | 0.89 | 0.98 |
|  | **high fever** | **0.77** | **0.77** | **1.0** |
|  | cough | 0.90 | 0.90 | 1.0 |
|  | **dry cough** | **0.68** | **0.68** | **0.93** |
|  | wet cough | 1.0 | 1.0 | 1.0 |
|  | congestion | 1.0 | 0.98 | 0.98 |
|  | **nasal congestion** | **1.0** | **0.97** | **0.97** |
|  | pain | 0.83 | 0.83 | 1.0 |
|  | **chest pain** | **0.98** | **0.99** | **0.99** |
|  | **muscle pain** | **0.60** | **0.60** | **0.98** |
|  | shortness of breath | 0.98 | 0.95 | 0.98 |
|  | **dyspnea** | **1.0** | **0.99** | **0.99** |
|  | tachypnea | 0.79 | 0.79 | 1.0 |
|  | **malaise** | **0.41** | **0.40** | **0.99** |
|  | headache | 0.68 | 0.65 | 0.97 |
|  | **sore throat** | **0.99** | **0.99** | **1.0** |
| **Findings** |  | 0.94 | 0.93 | 0.99 |
|  | **hypoxia** | **0.81** | **0.80** | **0.99** |
|  | opacities | 1.0 | 1.0 | 1.0 |
|  | ***bilateral opacities*** | ***0.85*** | ***0.80*** | ***0.96*** |
|  | infiltrates | 1.0 | 1.0 | 1.0 |
|  | ***lung infiltrates*** | ***1.0*** | ***0.99*** | ***0.99*** |
| **Disorders** |  | 0.94 | 0.93 | 0.99 |
|  | respiratory distress | 0.97 | 0.97 | 1.0 |
|  | ***acute respiratory distress syndrome*** | ***0.97*** | ***0.94*** | ***0.97*** |
|  | ARDS | 0.80 | 0.79 | 0.99 |
|  | ***pneumonia*** | ***1.0*** | ***1.0*** | ***1.0*** |

